# Individual reference intervals in practice: A guide to personalise clinical and omics level data with IRIS

**DOI:** 10.1101/2022.07.18.22277788

**Authors:** Murih Pusparum, Olivier Thas, Gökhan Ertaylan

## Abstract

Reference intervals (RI) are the best-established methodology used for the interpretation of numerical clinical level data in healthcare and clinical practice. As the test results are interpreted by comparing with the (population-derived) reference intervals, the quality of the calculation and implementation of reference intervals play a major role in decision-making process at the subject level. Here we describe the IRIS workflow to compute Individual Reference Intervals (IRI) based on multiple “healthy” data points from the same subjects and also utilising peers’ test results. We have improved the IRI models so they allow for covariate adjustments, such as sex and age. The IRI is expected to play pivotal roles in i) early detection of disease transition in chronic diseases by facilitating the detection of small deviations in clinical measurements, ii) monitoring personal disease progression, either using the standard clinical biochemistry test results or the omics level data. We demonstrate the utility of IRI in clinical and omics level data (proteomics and metabolomics) from two different longitudinal studies, including prior data processing and data quality check procedures. We have created an integrated application IRIS incorporating all described steps in an easy-to-use tool in research and/or clinical practice. We compute the IRI estimates in a healthy population to demonstrate its diagnostic utility in chronic diseases and from a diseased cohort to demonstrate its potential in disease monitoring.

## Introduction

Last decade has seen significant advancement of modern statistical and AI methodologies for interpreting numerical healthcare information, such as the utilisation of longitudinal health record data from an individual for the early disease detection [1]. However, we have not yet witnessed significant changes in routine healthcare and clinical practice partially due to the lack of statistical methodologies fit for purpose to handle these data at the personal level and the blackbox nature of some of proposed algorithms [2; 3]. It is still common practice today when consulting a medical practitioner, a biological sample is taken and some clinical laboratory tests are run testing for clinical biomarkers that are believed to be associated with potential disease(s). For any of these biomarkers, a reference interval is compared with the test result of that sample; when the result is within the interval, a normal reading is declared, otherwise further clinical tests may be advised.

The current reference interval estimation procedures mostly rely on cross-sectional data, i.e. a cross-sectional population of healthy subjects. This constraint makes them difficult to be implemented for early disease diagnosis, since patients at the early stage often behave similarly as the general healthy population. A novel concept of a personalised version of reference interval has been recently introduced; these intervals are referred to as the Individual Reference Intervals (IRI) [4; 5]. This idea enables a more precise interpretation of reference intervals as it utilises historical individual data, and hence the IRI is more personalised and narrower than the population-based version. This makes the IRI an appropriate tool for assisting the early stage disease diagnosis.

In the last few years, extensive efforts have been made for integrating multi-omics data in order to provide a better understanding at the molecular level of diseases, including chronic diseases [6; 7]. Unlike the clinical biochemistry data, omics technologies usually give vast information in a high-throughput and multi dimensional fashion. It is therefore considered cost effective and comprehensive [8]. Some omics technologies, such as proteomics and metabolomics, are now even frequently incorporated in routine biological and medical studies [9].

By definition, chronic diseases require medical attention that last for one year or more, and they may progress over time[10]. Patients with chronic diseases are frequently diagnosed late, when symptoms are set and the prognosis is poor. From this perspective, we hypothesise that the importance of utilizing omics data in defining disease sub-types and relationships between chronic diseases is self evident, the IRI approach is instrumental in the diagnosis of disease onset. In this manuscript, we aim to answer the question: **How can we utilize individual’s own data together with his/peers’ as baseline to calculate reference intervals for a key parameter(s) meaningful for identifying disease onset/progression**. We have developed and described an extension of the IRI method of Pusparum et al. [5], which allows for the adjustment of the IRI for covariates (e.g. age, sex). The methods are illustrated on clinical biochemistry and omics data (metabolomics and proteomics). In particular, two real-life datasets: the unique high-dimensional clinical and multi-omics longitudinal data of IAM Frontier study (IAF) [11] and the longitudinal multi-omics data of a case-control study in patients with irritable bowel syndrome (IBS) [12]. With both datasets we have performed an association analyses for discovering the relationships between several chronic diseases (CVD, CKD, and IBS) and omics data. For some selected clinical and omics biomarkers, the IRIs are computed. These procedures are further referred as the *IRIS workflow*, which includes a pipeline for automated checks for some of the assumptions underlying the IRI. The pipeline is available as an R Shiny app: IRIS. The procedures are explained in detail in Section 2. We employ our recent IRI estimation methodology and its extensions developed by Pusparum et al. [5] for calculating all IRIs in this study. We also show two crucial roles of IRI in **early detection of disease transition** and in **monitoring personal disease progression**.

Throughout this manuscript, 1) a *subject* has the same meaning as a person/an individual; 2) *subject time series* refer to a series of a clinical or omics values measured in one subject over a period of time; 3) a *feature* means a particular biological parameter that is measured from a laboratory test or from omics technologies i.e. a clinical feature comes from a clinical laboratory - total cholesterol, glucose, creatinine are clinical features while KIM-1 and CCL-28 are proteomics features; and 4) a *biomarker* refers to a particular biological feature which serve as indicator for health- and physiology-related assessments, or signs of a normal or abnormal process (related to diseases).

## Materials and methods

### Datasets

Two datasets were used: the IAM Frontier (IAF) study and a study on the irritable bowel syndrome (IBS). The IAF dataset comes from a unique small-scale, high-dimensional longitudinal cohort study that ran for 13 months in 30 healthy subjects, consisting of 15 male and 15 female participants. The study specifically targeted healthy subjects within the age range of 45-59. The subjects were selected based on the inclusion criteria of not suffering from a chronic disease, diagnosed and currently followed-up by a medical specialist, including asthma, chronic bronchitis, chronic obstructive pulmonary disease, emphysema, myocardial infarction, coronary heart disease (angina pectoris), other serious heart diseases, stroke (cerebral haemorrhage, cerebral thrombosis), diabetes, cancer (malignant tumour, also including leukaemia and lymphoma). The age range was selected because the highest prevalence of onset of these chronic diseases occurs from the age of 45-65. At monthly visits, after an overnight fasting, samples (whole blood, plasma, urine, stool) were collected and sent to accredited laboratories. Comprehensive multi-omics and clinical biochemistry data were assessed. Self-administered questionnaires on, for example, health conditions and physical activity were also completed by the participants. In this article we considered the clinical biochemistry, proteomics, and metabolomics features of the IAF study. The clinical biochemistry data consist of 88 clinical features that were monthly measured by an accredited clinical laboratory. Furthermore, bi-monthly data of 249 metabolites measured by NMR metabolomics analytical techniques and 266 proteins from OLINK cardiovascular and inflammation panels were included in our analyses. All subjects participated in the study have given their consent for displaying their anonymised pseudo-names for research purposes i.e. we do not disclose any of the participants’ real names.

The IBS data were collected from a longitudinal case-control study that observed and compared healthy subjects to two types of IBS patients: IBS-C (constipation-predominant) and IBS-D (diarrhoea predominant)[12]. In this study, a total of 77 participants were asked to donate their stool samples on a monthly basis until the sixth month. However, not all of them provided full samples; therefore, we only included participants with at least three time points measurements. Monthly metagenomic sequencing and NMR metabolomics were also assessed, together with a dietary survey of food intake and symptom severity at each visit. For this article, the longitudinal NMR metabolomics dataset consisting of 24 metabolites are analysed. Table 1 presents a further description of both datasets.

**Table 1.** Overview of the two datasets used in this paper (IAF and IBS)

| Data | Cohort | M* (n) | F* (n) | Age (years) | #features |
| --- | --- | --- | --- | --- | --- |
| IAF | Healthy<sup>†</sup> | 15 | 15 | 45-59 (4.24) | 88 clinical<br>249 metabolites<br>266 proteins |
| IBS | Healthy<sup>†</sup> | 5 | 19 | 23-59 (10.89) | 24 metabolites |
|  | IBS-C | 1 | 21 | 20-63 (14.21) |  |
|  | IBS-D | 10 | 19 | 21-61 (12.40) |  |

<sup>†</sup> Free of chronic diseases at baseline.
\* M is male, F is female participants.

### IRIS workflow

The IRIS workflow consists of two steps: IRIS feature selection and IRIS pipeline. In the first step, a set of features is associated with the target disease phenotype of interest and a few of the most relevant features is selected. For the implementation in preventive practice, we encourage to use phenotype in a form of predictive risk scores and therefore an IRI would serve as a predictive tool to assist an early disease detection. In the case of when the diagnosis has been carried out w.r.t. a particular disease, the set of features may still be associated with the binary outcome resulted from the diagnosis (1 if disease is diagnosed and 0 otherwise). The IRI would then allow to monitor the disease progression based on the relevant selected features. In the second step, the selected features, we further refer them as *biomarkers*, are entered the IRIS pipeline. In this pipeline, we analyse each biomarker following the assumption that the subject time series should be in a stable state. Still in the pipeline, we finally compute the IRI for each biomarker.

### IRIS feature selection

The aim of IRIS feature selection step is to identify (potentially) relevant features with the phenotype of interest. We have included two datasets to demonstrate unique implementations of IRIS workflow in: IAF data to demonstrate ability in the early disease detection which needs a risk distribution; IBS data on identifying potentially disease progression and parameters to look across disease sub-phenotype.

For the IAF dataset, clinical data were collected in a high throughput fashion (as per cohort protocol) via laboratory tests where in practice, usually are available from individuals who are tested to assess the risks of abnormalities or (chronic) disease onset. Many clinical features measured in laboratory tests are known to be associated with an illness or a chronic disease, such as an abnormal level of albumin may indicate liver diseases, high creatinine values can be a signal of kidney failures, or blood glucose levels that are used to identify the diabetes status. Hence, an IRI can also be estimated for any clinical biomarkers, for detecting abnormalities that may associate with a (chronic) disease.

The IAF clinical data is used to compute a 10-year cardiovascular disease (CVD) risk score [13], with the following covariates included in the model: age, sex, race, smoking status (yes/no), systolic blood pressure, diabetes (yes/no), HDL cholesterol, total cholesterol, and treatment for hypertension (yes/no). We further performed an association analysis between the CVD risk scores and each set of metabolomics and proteomics data. A simultaneous penalised linear mixed model (SP-LMM), as implemented in the *splmm* R package, was fitted for each dataset. This involves a feature selection in a high-dimensional longitudinal setting [14]. A few metabolites and proteins with the largest absolute effect size were considered to be potentially associated with the CVD onset. These biomarkers were then selected and included in the IRIS pipeline. The detailed workflow is depicted in Figure 1A.

**Fig. 1.**
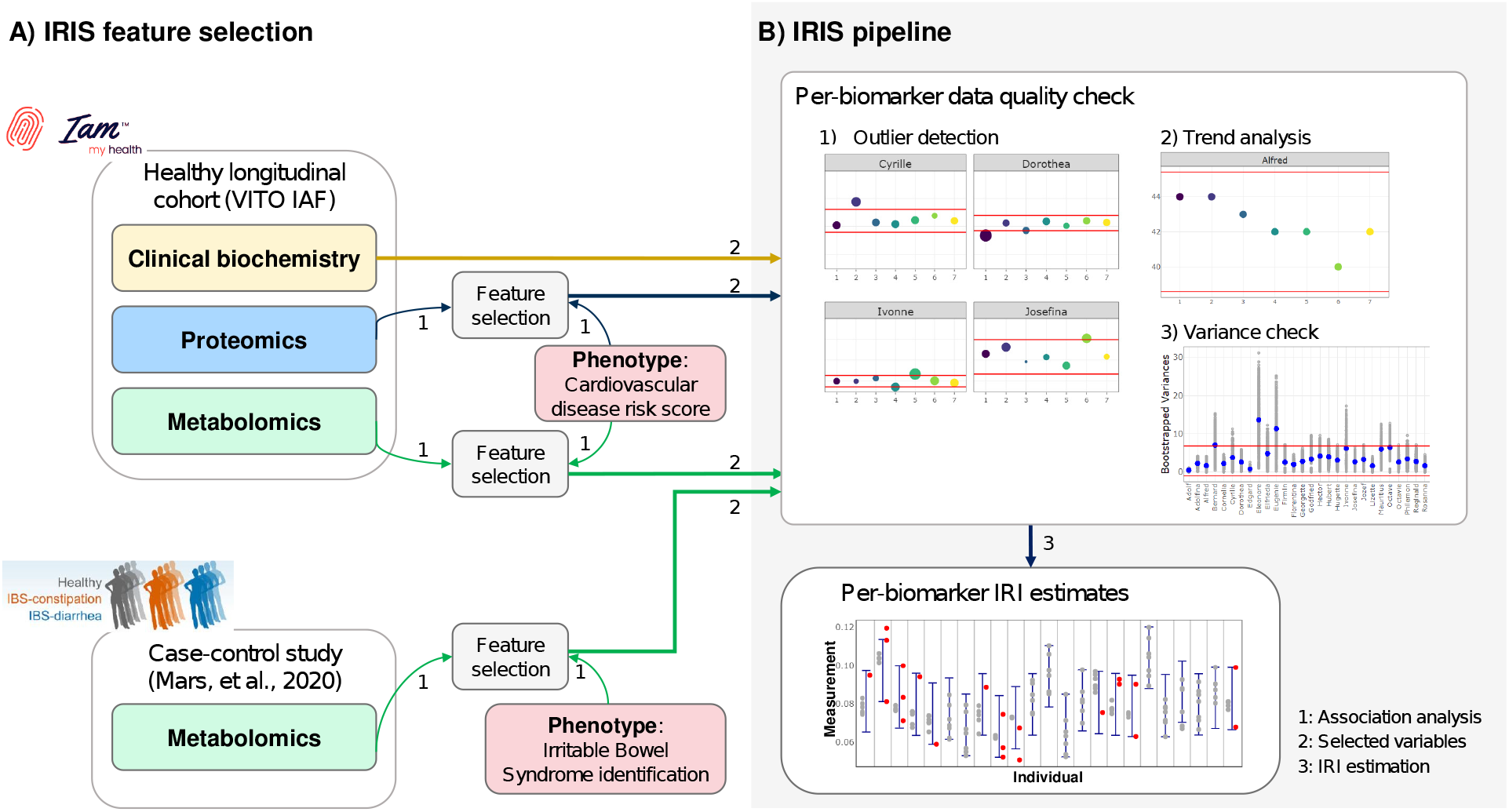
IRIS workflow for feature selection and IRI calculation. (A) Two clinical and omics datasets are used in this study (IAF and IBS). In the IRIS feature selection, association analyses are performed on each omics dataset, resulting in a set of potential biomarkers. (B) The selected biomarkers enter the IRIS pipeline; this includes a data quality check before the calculation of the IRI for each combination of a biomarker and a subject.

For the IBS data, an association analysis is performed for selecting features that can differentiate a healthy subject from an IBS patient. A generalized linear mixed model with *ℓ*_1_ penalisation was fitted on the log10-transformed IBS metabolomics data and metabolites with the largest absolute effect size were selected. A binary response of subjects’ status (healthy and IBS patient) was considered. For fitting the model, we used the implementation of the *glmmLasso* R package [15]. Standard *t*-tests with a Benjamini-Hochberg (BH) correction are carried out for comparing the IRI widths between the healthy subjects and IBS patients. In both the IAF and IBS modelling, the Bayesian Information Criterion (BIC) value is used for the model selection.

### IRIS pipeline

For each selected biomarker, we performed a data quality check for ensuring that the subject time series are in a stable state i.e. the subjects are in a healthy condition over a period of time. This procedure is part of the IRIS pipeline (Figure 1B) and it has been implemented in the IRIS application. The pipeline starts with checking if there are outlying observations in each time series of one subject. Outliers are defined in terms of the median absolute deviation (MAD). For each time series, the MAD threshold was computed. In particular, 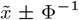(0.99)MAD was used to set the lower and the upper thresholds for the calling of outliers, where 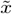 is the median of each subject time series and Φ^*−*1^(0.99) is the 99^*th*^ quantile of a standard normal distribution. We also computed the thresholds and defined outliers for all other features in the data (besides the selected biomarkers). For each time point in every subjects, the proportion was taken i.e. the number of features with outliers proportional to the total number of features. By default, all outliers will still be included in the IRI estimation, as they might contain important information and professionals’ opinions are required to do otherwise. However, for this study, an arbitrary threshold of 20% was set i.e. if a particular measurement in the time series is also outlying in more than 20% of the total features, we assume abnormalities and hence that measurement will be excluded. All outliers that are still included and will further be flagged in the final IRI estimates.

A stable state of subject time series was then assessed using the nonparametric Mann-Kendall (MK) test of monotonic trends [16; 17]. We argue that if a monotonic trend is present within one each time series, it may suggest a decline or a progression towards a particular health condition (disease), hence an unstable condition. We chose a nonparametric test, because in realistic datasets each subject contributes only short time series (i.e. small sample size). Apart from the MK test for analysing if either a monotonic increasing or decreasing trend is present, for each subject, we also computed Spearman rank correlation coefficients between subject time series and the time point covariate. In case a monotonic trend and the correlation are statistically significant, the subject will be excluded from the IRI estimation. The last step in the data quality check involves the computation of the variance in each time series. To support the argument that the subject time series should be in a stable state, the variance of each time series should also remain small for all subjects. A similar MAD threshold was computed; subjects with variance exceeding the threshold will be excluded. The pipeline overview can be consulted in the *Supplementary Document* Figure S1.

### Overview of IRI methods

We have implemented the Penalised Joint Quantile Model 2 (PJQM2) for simultaneously estimating the lower and the upper bounds of the IRIs [5]. This method does not rely on distributional assumptions, but it makes use of quantile models that inherit the typical flexibility of the statistical models. With *τ*_1_ (here *τ*_1_ = 0.025) and *τ*_2_ (here *τ*_2_ = 0.975) representing the probabilities corresponding to the upper and the lower bounds of the IRIs, the corresponding quantiles are modelled as:

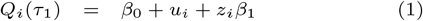

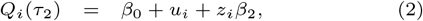

where *β*_0_ is the fixed intercept and *u*_*i*_, *z*_*i*_ are the subject-specific effects. Further, *β*_1_ and *β*_2_ are the parameters that allow for the subject-specific IRI widths. The model contains two subject-specific random effects, *u*_*i*_ and *z*_*i*_, that allow for between-subject variability. A parameter estimation procedure involving an *ℓ*_2_ penalty terms is described in details in Pusparum et al. [5].

The model allows for covariate adjustment by simply including additional terms for any covariates. If, for example, age affects the outcome, then better IRIs can be constructed if information of subjects of the same age can be shared; this is approximately the result of these extended models. As the original theoretical IRI implementation of Pusparum et al. [5] does not allow for it, as part of this paper, we have extended our earlier PJQM2 implementation so that covariates can be included in practice.

In our examples, sex and age are included with the reasoning subjects’ genetic and biological make ups are generally different between males and females, and their clinical as well as molecular level measurements may change as part of the aging process. The IRI model now become

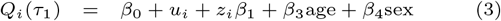

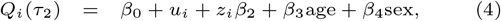

where *β*_3_ and *β*_4_ are the parameters for the age and sex effects. The parameter estimation procedure essentially remains the same as in (1) and (2), except that now Linear Quantile Mixed Model (LQMM) [18; 19] are fitted with age and sex as covariates.

Recently, Coşkun et al. [20] proposed another method for the calculation of a personalised version of reference intervals. However, their technique relies on the normal distribution. In real-life settings, it is often difficult to check a distributional assumption, particularly in rather short-time series such as in the IAF and IBS datasets. For these reasons, we have not used this method in this study.

## Results

### IRI in clinical biochemistry as a standard tool for assisting diagnosis in clinical laboratory test

We aimed to estimate IRIs for some clinical biochemistry features available the IAF longitudinal dataset. In particular, we decided to estimate IRIs of creatinine, as it is widely known that high levels of creatinine in the blood correspond to kidney failures, leading to chronic kidney diseases (CKD). In the IAF study, we collected monthly samples for clinical laboratory tests over a one year period. However, we only used the first seven time points for estimating the IRIs and took the last creatinine measurement at time point 12 or 13 (after *±*6 months after the last measurement) for demonstrating the interpretation of the IRIs.

Before estimating the IRIs, as part of the IRIS pipeline, we have first checked for outliers, the presence of monotonic trends, and the variance of each subject time series. We observed that several subjects had outlying observations, but we did not detect any subjects with outliers in more than 20% of the clinical features. The trend analysis and the variance checking showed that one subject showed an increasing monotonic trend, and two subjects had obvious distinct variances as compared to their peers. Therefore, these three subjects were excluded from the IRI estimation. Further details on this data quality step can be consulted in the *Supplementary Document* Figures S2-S4.

Figure 2 shows the IRI estimates of the creatinine level, computed using PJQM2 method, with age and sex as additional covariates. As expected, the IRIs are stretched around each subject time series and the locations as well as the widths vary between the subjects. We also see, for example for Adolfina, that the upper bound is expanded farther away from her time series measurements. This is in fact a desirable consequence of the between-subject information-sharing property of the method[5]: Adolfina has overall smaller observations than the other subjects, and this is reflected in her IRI, which adapts to Adolfina’s own data, but also expands into the direction of the bulk of the data.

**Fig. 2.**
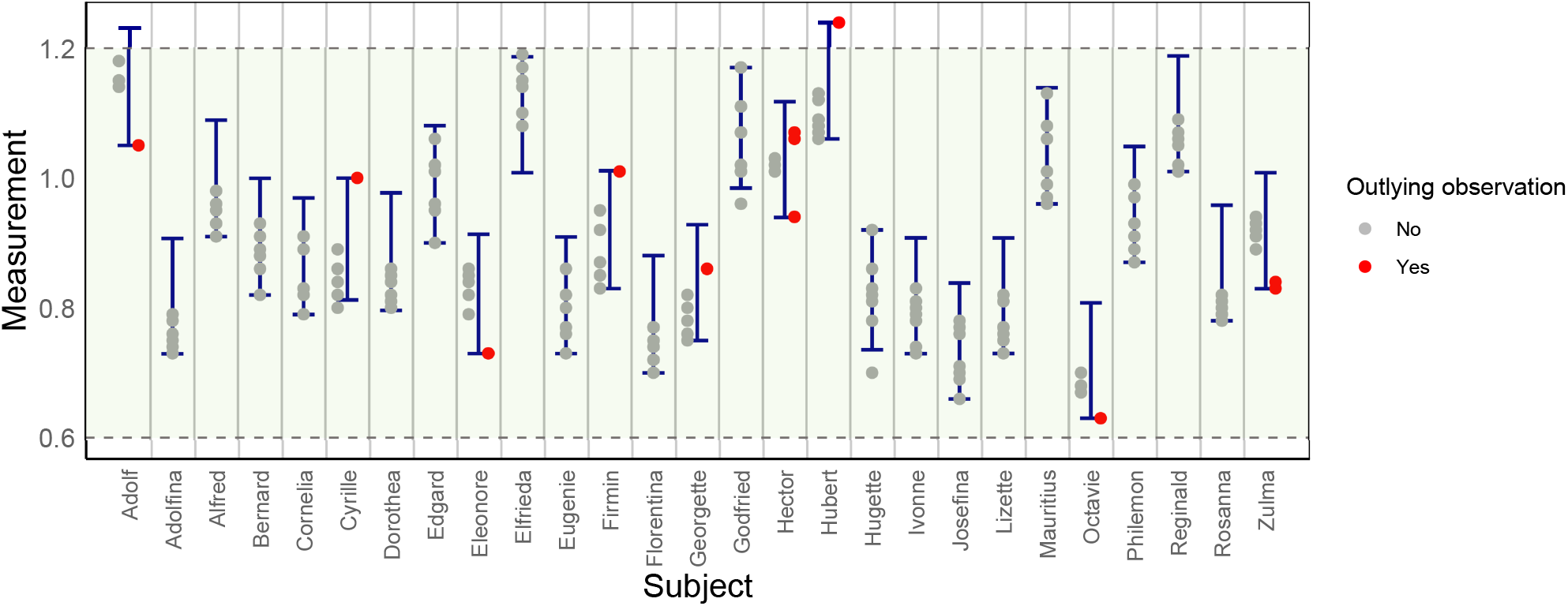
The estimated IRIs of creatinine (mg/dl) in the IAF clinical biochemistry data, with age and sex as additional covariates in the model. The circle dots refer to the subject time series used for the IRI estimation. Outlying observations are included and they are flagged and showed as red dots.

The IRIs in practice is shown in Figure 3, where the historical data (used for estimating the IRIs) and new measurements are plotted together, with corresponging IRIs. From the results we can see that the future measurements of Alfred, Florentina, and Hubert are below the lower IRI bound, while for Adolfina, Rosanna, and Zulma they are somewhat on the borderline. For these subjects, their last measurements can be an early sign of hypocreatinemia resulted from the failures of liver function and low-protein diet Had those new measurements been located in upper borderline or outside the upper bound, it might have been an early caution of CKD onset.

**Fig. 3.**
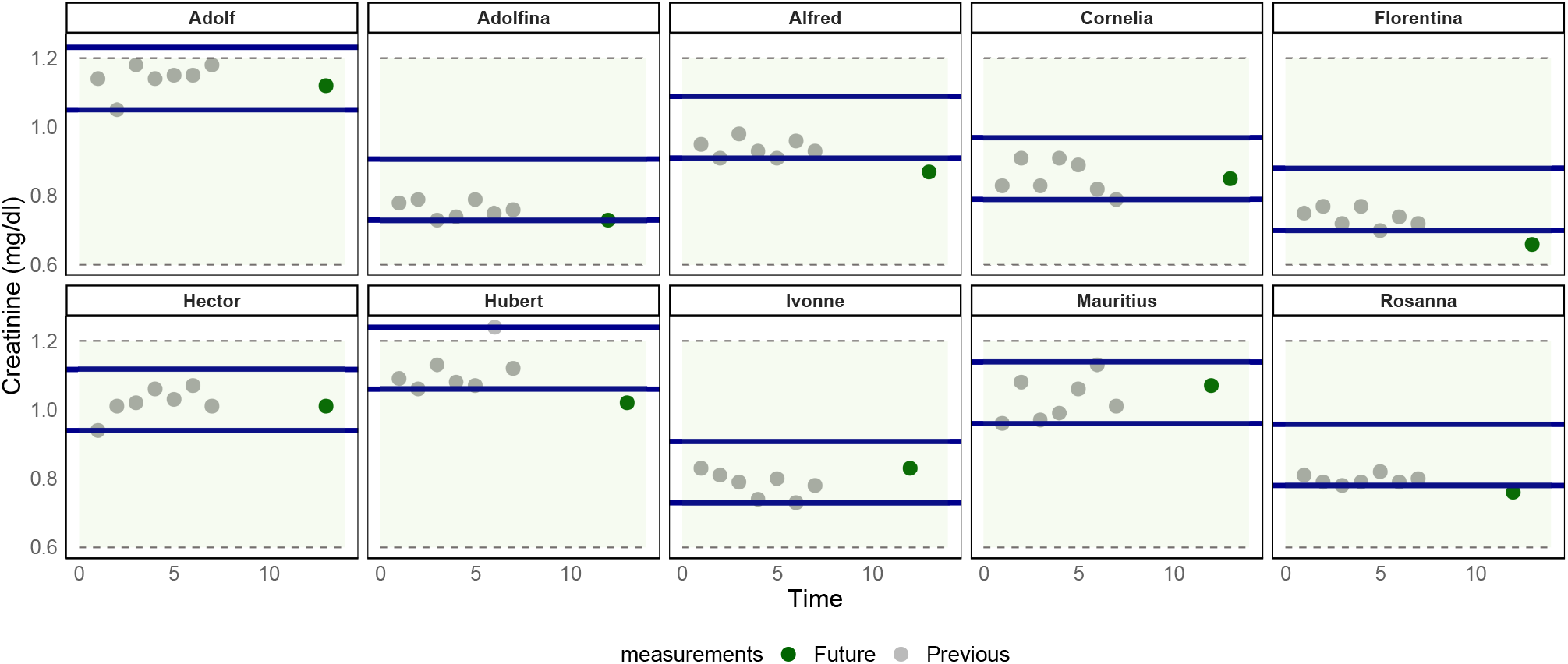
The estimated IRIs of creatinine (mg/dl) of ten subjects in the IAF clinical biochemistry data. Seven previous measurements (grey circles) were used for the estimation. The blue horizontal lines represent the IRIs, the shaded green areas are the population-based reference intervals [21]. A green dot is presented for each subject referring to a future measurement i.e. the last measurements collected in the study and not used for the IRI estimation. The IRIs should be used to interpret the future measurements. E.g. the new measurement of Hubert suggests a creatinine abnormality.

### IRIs derived from omics data provide personalised normal values for CVD diagnosis

Clinical laboratory tests are often considered as a golden standard in assisting disease diagnosis. Alternatively, we have also seen the prospective of omics data such as proteomics and metabolomics in supplementing medical diagnosis, as they are likely to be close to phenotype and therefore may benefit as a disease’s biomarker [6]. In addition, omics technologies are also becoming cheaper and more accessible [6], offering a more affordable analysis to complement the common clinical laboratory test. In this study, using the IAF data we examined the most predictive metabolites and proteins w.r.t. cardiovascular diseases (CVD). The IAF data was indeed collected from ‘apparently’ healthy subjects who did not suffer or were not diagnosed with CVD. However, as the participants were in the highest prevalence state of chronic disease onset, according to their age, there is still a risk related to lifestyles and physical activities. From the IRIS feature selection, we identified five and ten most discriminating metabolites and proteins related to the 10-year CVD risk scores. The *Supplementary Document* Section 1.1 can be consulted for further details of the model. Table 2 explains the list of selected proteins and metabolites together with the estimated effect size. In this manuscript, for illustration purposes we chose citrate and phospholipids in small density LDL cholesterol (S-LDL-PL) from the metabolomics dataset as they give the largest effect size. Analogously, ICAM-2, SELP, and KIM1 were chosen from the proteomics dataset. The scatter plots of these biomarkers against the CVD risk scores in Figure 4 shows a negative association between the CVD risk scores and citrate as well as the ICAM-2 protein, consistent with their negative estimated effect sizes.

**Table 2.** Estimated effect sizes of each metabolites and proteins from the metabolomics and proteomics association analysis against CVD risk scores. The selected biomarkers are in bold face.

| Metabolomics |  | Proteomics |  |
| --- | --- | --- | --- |
| Biomarker | Estimate | Biomarker | Estimate |
| (Intercept) | -24.1155 | (Intercept) | -28.0100 |
| Age | 0.3913 | Age | 0.0266 |
| sex | 5.7284 | sex | 1.2833 |
| **Citrate** | **-5.2821** | CCL28 | 3.0866 |
| Glucose | 0.3252 | GRN | 1.6412 |
| Linoleic acid | 0.5356 | **ICAM-2** | **-11.8859** |
| Lactate | 0.2132 | IL2-RA | 3.3319 |
| **S_LDL_PL** | **20.2414** | **KIM1** | **3.3424** |
|  |  | MARCO | 0.2771 |
|  |  | MMP-10 | -0.7601 |
|  |  | **SELP** | **6.0702** |
|  |  | SHPS-1 | -2.8197 |
|  |  | THPO | -0.2302 |

**Fig. 4.**
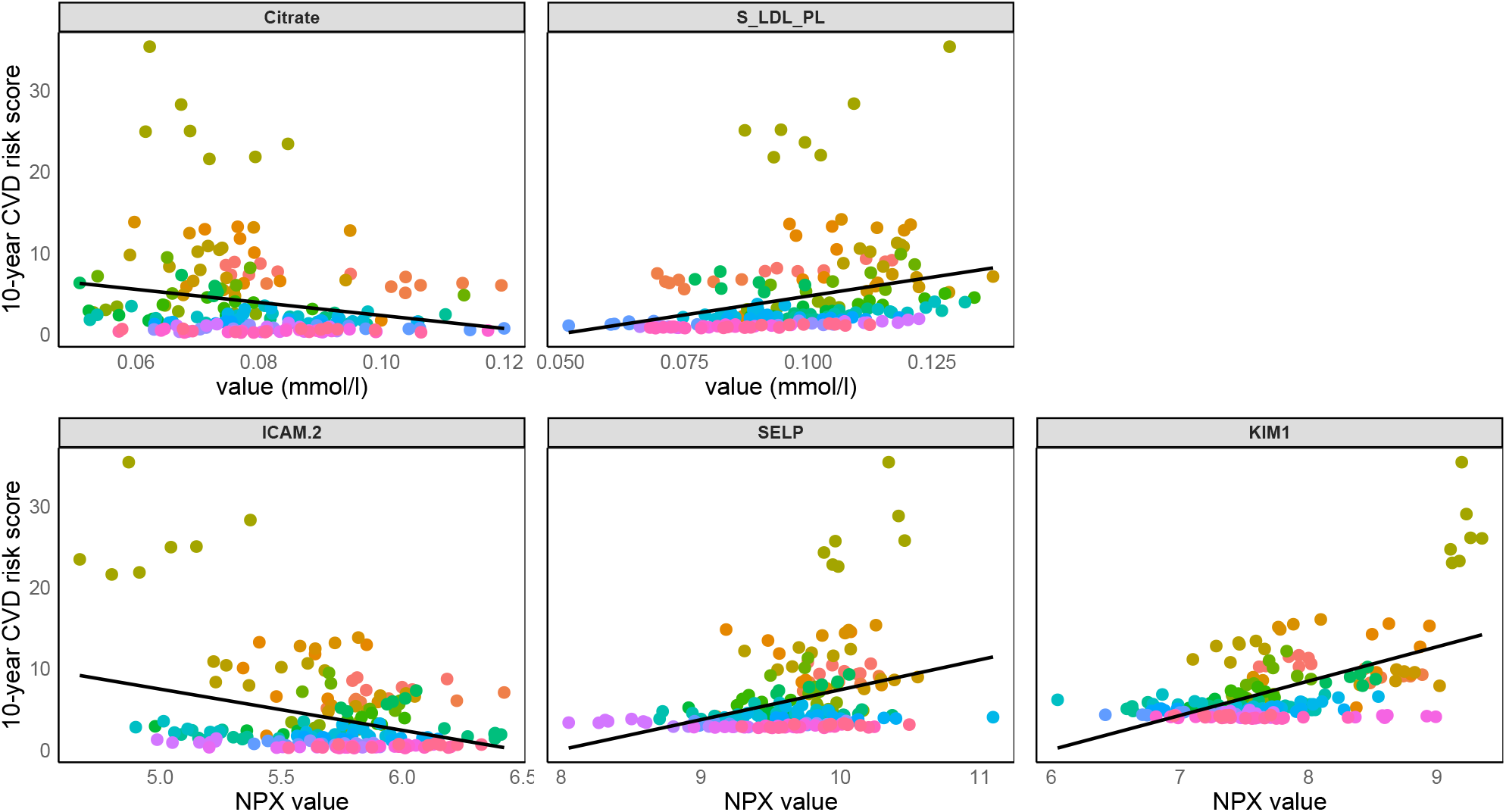
Scatter plots of the selected biomarkers in the metabolomics and proteomics datasets with their corresponding regression lines (black solid lines). Each dot refers to a measurement at one time point; different colors are assigned to different subjects. High citrate and ICAM-2 abundances suggest lower 10-year CVD risk scores.

We performed a data quality check as explained in the IRIS pipeline in Figure 1B for each metabolite and protein. Figure 5 presents the complete outlier identification of citrate. For this metabolite, we found that Octave’s citrate measurement at the first time point is outlying, and at the same time point, his measurements are also outlying in 21.3% metabolites. Therefore, we excluded his first citrate measurement from the data. The second measurement of Cyrille should also be removed (26.9% of metabolites are outlying at this time point), but at the end, we completely excluded Cyrille from the estimation as a monotonic trend was significantly present. Based on the trend analysis and variance checking (see *Supplementary Document* Figure S5-S6), seven subjects were eventually excluded.

**Fig. 5.**
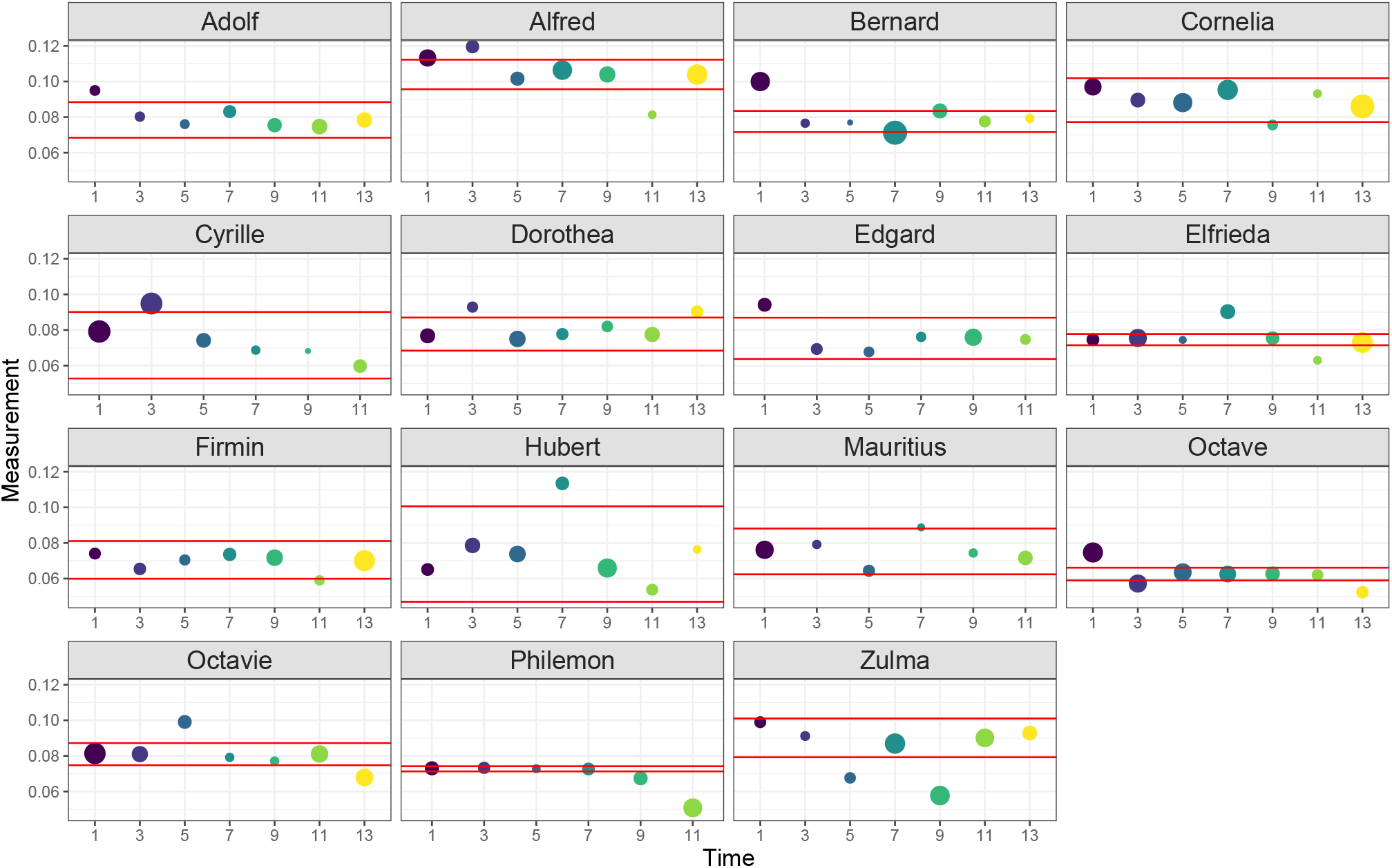
Outlier analysis step for citrate in the IAF metabolomics data. Outlying observations are shown as the circles outside the MAD thresholds (red horizontal lines). An outlying observation is removed if the observation at the same time point is also outlying in more than 20% of the features in the dataset. An example: Cyrille’s measurement at the second time point.

Figure 6 shows the estimated IRIs of citrate in metabolomics dataset, with age and sex as additional covariates in the model. Similarly, these IRIs can be used for interpreting the future measurements of the subjects. From Table 2, a negative coefficient was estimated for citrate, meaning that higher citrate levels are more favourable w.r.t. CVD. The IRI interpretation may hence be more focused on the future measurements below the lower bound. Results of other selected metabolites and proteins can be found in the *Supplementary Document* Section 3.1.

**Fig. 6.**
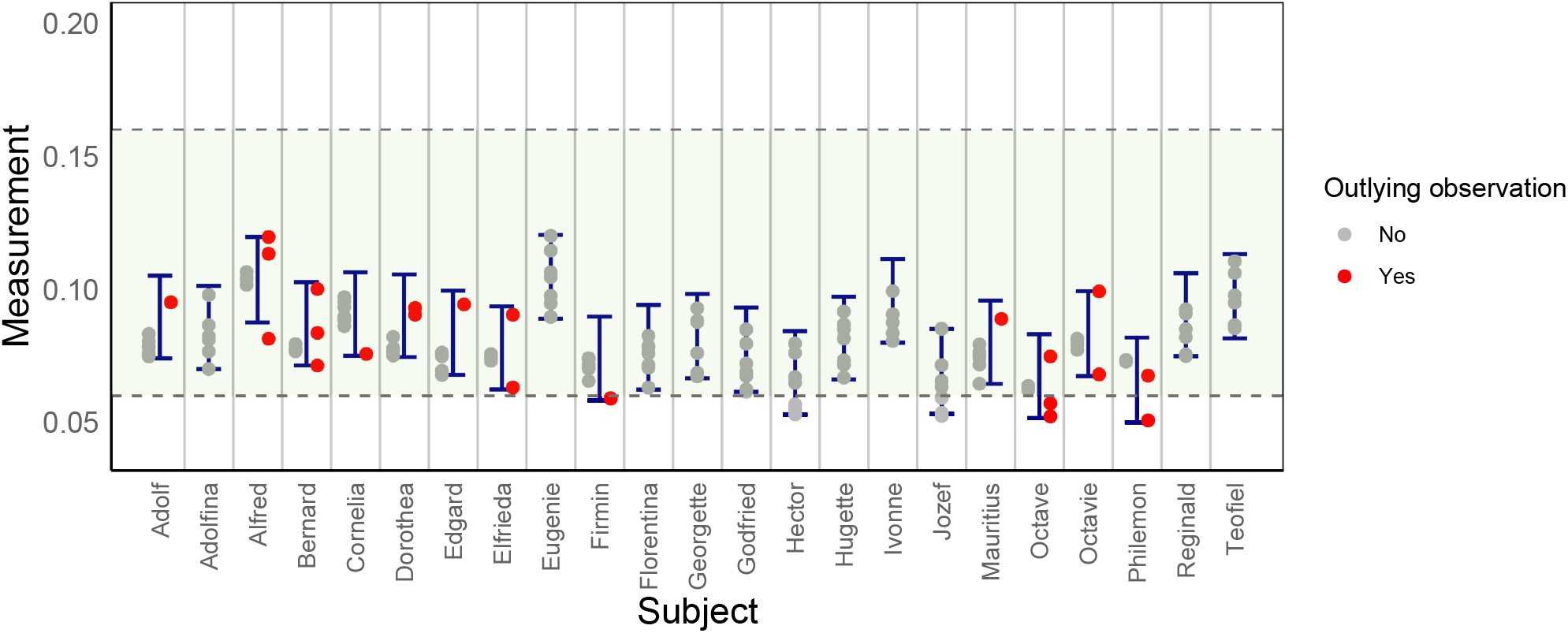
The estimated IRIs of citrate (in mmol/l) in the IAF metabolomics data, with age and sex as additional covariates in the model. The circle dots refer to the subject time series used in the estimation. Outlying observations are included and they are flagged and shown as red dots.

### IRI as a monitoring tool for subjects with Irritable Bowel Syndrome (IBS)

For the IBS dataset, we aimed to estimate IRIs for biomarkers measured from both healthy and diseased individuals diagnosed with IBS. Unlike the routine reference intervals that provide normal values for detecting abnormalities, here we show how we can utilise IRIs for disease monitoring. The IRIS feature selection suggested the three most discriminating metabolites: hypoxanthine, glucose, and b-arabinose (see *Supplementary Document* Section 1.2 for the model description). The effect size estimates as well as the estimated odds ratio are presented in Table 3. In this manuscript, we decided to show the analysis of hypoxanthine as the largest effect size is contributed by this metabolite.

**Table 3.**
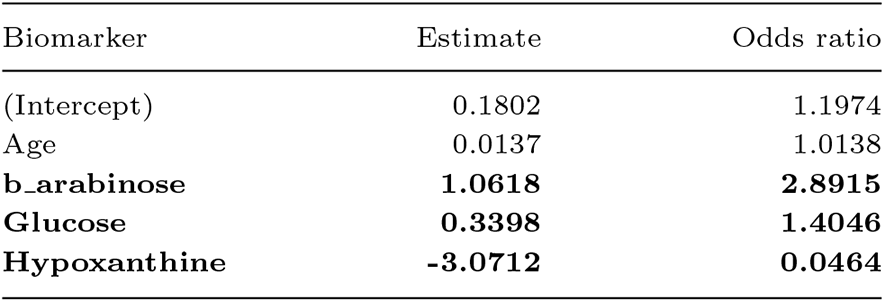
Estimated effect sizes of each biomarker from the IBS metabolomics association analysis against the IBS status. The selected biomarkers are in bold face.

| Biomarker | Estimate | Odds ratio |
| --- | --- | --- |
| (Intercept) | 0.1802 | 1.1974 |
| Age | 0.0137 | 1.0138 |
| **b_arabinose** | **1.0618** | **2.8915** |
| **Glucose** | **0.3398** | **1.4046** |
| **Hypoxanthine** | **-3.0712** | **0.0464** |

Figure 7 presents the estimated IRIs for hypoxanthine for the three types of cohorts, after the data quality check was done in the IRIS pipeline. For the healthy and IBS-D IRIs, the estimates were obtained with models that include both age and sex. For the IBS-C IRIs, however, we only incorporated age (due to a highly imbalance of sex covariate). We clearly see that healthy subjects in general have larger hypoxanthine IRIs, both in terms of location as in terms of width, than the IBS patients. It has been shown that fecal hypoxanthine abundances were significantly lower in IBS-C and IBS-D patients, and therefore the decline of hypoxanthine abundances has implication to the IBS pathogenesis [12]. Similarly, we also observed that the IRI widths are clearly smaller in IBS-C and IBS-D patients, as compared to the healthy subjects (see *Supplementary Document* Table S1).

**Fig. 7.**
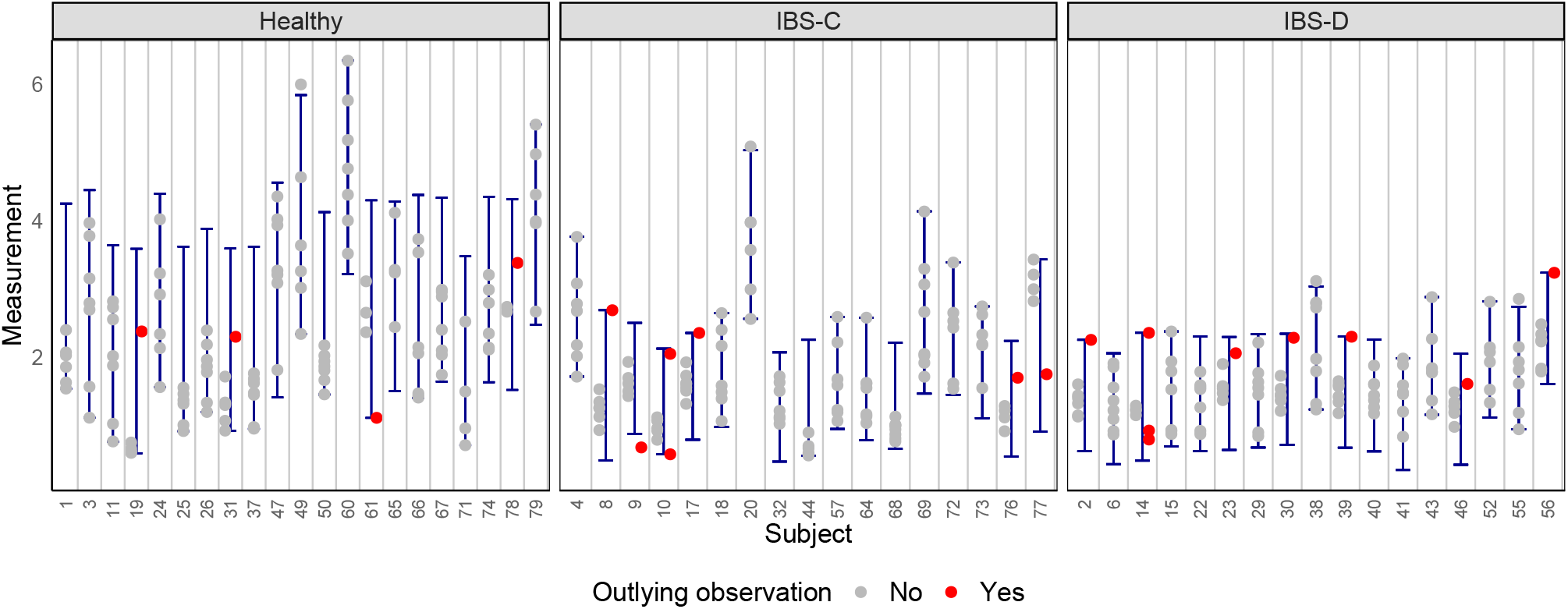
The estimated IRIs of hypoxanthine (relative abundance) in the IBS metabolomics data for the healthy cohort, IBS-C and IBS-D patients.

The healthy IRIs can be interpreted as the normal values, where if these subjects have new measurements outside their IRIs, especially below the lower bound, it should be an early warning of a potential IBS disease. The interpretation becomes slightly different for the IBS IRIs. For IBS patients, the estimated IRIs can serve as a monitoring tool for disease progression, either an improvement or a decline, as a result of e.g. an intervention from drug therapy or medication. To illustrate, if the next hypoxanthine measurement of subject 2 in the IBS-D group is larger by 1 unit than his/her IRI, it then would suggest an improvement. Certainly, this analysis should also be still supported by the professional assessments. Several IRIs from the IBS-C group appear to deviate from their peers, e.g. IRIs of subject 4, 20, and 69. These IRIs are similar in location and width as the healthy IRIs, and they perfectly capture the within-subject variation. If the individuals in this group are truly IBS-C patients, these IRIs demonstrate that the IBS state can differ from one subject to another w.r.t. to hypoxanthine. Results of other selected metabolites can be consulted in the *Supplementary Document* Section 3.2.

## Discussion

Here we propose IRIS as a workflow for analysing clinical and omics data to complement early-stage diagnosis and monitoring of chronic diseases. The IRIS workflow consists of two steps: IRIS feature selection and IRIS pipeline. In IRIS feature selection, we describe procedures that can find the most discriminating features in clinical, metabolomics and proteomics datasets, w.r.t. cardiovascular diseases (CVD) and irritable bowel syndrome (IBS) as examples. The selected biomarkers are then analysed by computing Individual Reference Intervals (IRIs) [5; 4] with an adjustment of sex and age as the additional covariates. Prior to the IRI estimation, the IRIS pipeline that includes a data quality check is also proposed for checking the underlying IRI assumptions; the subject time series come from a healthy population (or a corresponding diseased population) in a stable state. An online IRIS application has been created incorporating all the steps in the pipeline for the research and/or clinical practice purposes. The workflow was implemented in clinical, metabolomics, and proteomics datasets collected in two longitudinal studies [11; 12].

In the clinical data, we show that the estimated IRIs of creatinine can support the diagnosis of CKD. Creatinine is commonly involved in the detection of CKD, particularly by calculating the estimated glomerular filtration rate (eGFR) which incorporates patients’ age and sex information [22; 23]. Similarly, the extension of the IRI estimation method to allow for covariates in the model (here: age and sex), makes it possible to complement the CKD diagnosis. In the IAF dataset, the IRI interpretation is not straightforward as all the samples were collected from healthy subjects (who did not suffer from CKD) in a one year period. Therefore, as an illustration, we took their last measurements and compared them to the IRIs estimated from the first seven time points. There are no subjects whose last measurements exceed the upper bound of IRIs, suggesting that the population was probably free from CKD during the course of the study. However, when a study follow-up is to be conducted, we can still use these estimated IRIs, as the models include age and sex of the participants.

In the metabolomics and proteomics IAF dataset, several parameters have been found to be associated with the 10-year CVD risk scores. Citrate and phospholipids in small LDL cholesterol (S-LDL-PL) are the most influential metabolites, and they have been previously found to be associated with CVD prediction [24; 25]. A negative association of citrate is supported by a finding that citrate has a protective effect during ischemia-reperfusion (I/R) injury that leads into morbidity or mortality associated with CVD [24]. The most discriminating proteins; ICAM2 and KIM1 have also been proved to be differentially expressed in coronary artery disease [26; 27]. An analogous interpretation as in the creatinine IRIs can be drawn from the metabolomics and proteomics IRIs. Since the metabolomics and proteomics data only consists of maximum seven time points, we did not show a similar implementation as in the creatinine IRIs.

The estimated IRIs can serve as a monitor for disease progression, especially for chronic diseases. We show the IRI estimates in hypoxanthine, where this metabolite was found to be associated with IBS. Increased level of fecal hypoxanthine abundances proved to improve the IBS progression, especially in IBS diarrhea predominant (IBS-D) patients [12]. Therefore, when fecal metabolomics measurements are available, the IBS patients can have their hypoxanthine abundances measured from time to time in order to monitor their IBS condition via IRIS.

Future studies with real-life clinical and omics data with different time series will give multitude of examples of the IRI implementation. The PJQM2 estimation method has a property of calculating IRIs of one single clinical or omics feature, taking into account subjects’ age and sex information. Integration with other related features or even other omics data could give a more comprehensive insight and would be immensely beneficial for the precision health domain.

**Key points**

- Individual reference intervals (IRI) give personalised interpretation of numerical clinical and omics data by utilising time series values from the same subject as well as peer’s results. They have unprecedented application potential in healthcare and clinical practice.
- We have expanded the state-of-the-art IRI model to allow for subject’s age and sex adjustment, and we share the guidelines in the IRIS workflow.
- We demonstrate the utility of IRI in clinical and omics data, both in healthy and diseased individuals, that can play a major role in the early detection of chronic diseases and personal monitoring of disease progression.
- We present an easy-to-use two step workflow packaged as a toolbox IRIS that integrates all the required steps prior to computing the IRIs.

## Data Availability

All data produced in the present study are available upon reasonable request to the authors.

https://data.mendeley.com/datasets/29n2z5r5ph/3

## Author contributions statement

M.P, O.T. and G.E. conceived the study, M.P. and G.E. designed and developed the IRIS workflow, M.P. and O.T. developed and improved the IRI methods, M.P. performed computational analyses, M.P, O.T. and G.E. wrote and reviewed the manuscript.

