## Supplementary Document for "Individual reference intervals in practice: A guide to personalise clinical and omics level data with IRIS"

### 1 Variable Selection for Omics Data

#### 1.1 Metabolomics and proteomics in the IAF data

The IAM Frontier (IAF) data consist of clinical biochemistry, physiological, and multi-omics measurements collected from samples of 30 'apparently' healthy individuals. Although the participants were considered healthy at the time of recruitment, the age of the participants were at the highest prevalence of chronic diseases (45-59 years old). Thus, 10-year cardiovascular disease (CVD) risk scores were computed, using the clinical biochemistry and physiological data, with the atherosclerosis cardiovascular diseases (ASCVD) calculator (see Appendix 7 in **?** ]). Variables included the risk score calculation are: age, sex, race, smoking status (yes/no), systolic blood pressure, diabetes (yes/no), HDL cholesterol, total cholesterol, and treatment for hypertension (yes/no). From the results, the male participants have 1.1-35.4% risk of CVD event(s) in the next 10 year, whereas the risk is only 0.3-3.9% in females.

These risk scores were calculated for each individual at each time point, and later will be utilised in the variable selection step of the data processing workflow. We associated both metabolomics and proteomics datasets (in two separate models) with the estimated CVD risk scores. We applied the simultaneous penalised linear mixed models (SP-LMM) implemented in *splmm* R package to perform a simultaneous variable selection of both fixed and random effects using a class of penalty functions as explained in **?** ]. This method was particularly developed for the purpose of variable selection in high-dimensional data. The following model was fitted for each metabolomics and proteomics data:

$$\mathbf{y}_i = \beta_0^{(n)} + \beta_1^{(n)}\text{age}_{ij} + \beta_2^{(n)}\text{sex}_i + \sum_{k=3}^{p}\beta_k\mathbf{X}_{ki} + b_0 + b_1^{(n)}\text{age}_{ij} + b_2\text{sex}_i + \epsilon_i,$$

where for the $i$-th individual with $m_i$ repeated measurements, $\mathbf{y}_i$ is the matrix of estimated ASCVD risk scores, $\mathbf{y}_i = (y_{i1}, y_{i2}, \ldots, y_{im_i})^T$ and $\mathbf{X}_{ki}$ is the matrix of fixed effects of the

$k$-th protein/metabolite covariates, $\mathbf{X}_{ki} = (X_{ki1}, X_{ki2}, \ldots, X_{kim_i})^T$. Fixed and random effect coefficients are denoted by $\beta_k, k = 1, 2, \ldots, p$, and $(b_0, b_1, b_2)$, respectively. The symbol $(n)$ indicates that for these parameters we keep them unpenalized. Two penalisation techniques are considered: LASSO and Smoothy Clipped Absolute Deviation (SCAD). The best model with the lowest Bayesian Information Criterion (BIC) was selected.

#### 1.2 Metabolomics in the IBS data

The variable selection procedure was also applied for the NMR metabolomics measurements in the IBS data. Generalised linear mixed models with $\ell_1$ penalisation were fitted to the log10-transformed metabolomics measurements and a binary response of IBS status. The following model was considered:

$$\text{logit}(\pi(x)) = \ln\left(\frac{\pi(x)}{1-\pi(x)}\right) = \beta_0 + \beta_1 \text{age}_{ij} + \beta_2 \text{sex}_i + \sum_{k=3}^{p} \beta_k \mathbf{X}_{ki} + b_0 + b_1 \text{age}_{ij},$$

where $\pi(x)$ is the probability of an individual is diagnosed with IBS, given the linear combination of individual's age, sex, and the set of metabolites. We performed a penalisation procedure implemented in the *glmmLasso* R package for estimating the effect sizes and selecting the most discriminating variables. Again, the best model with the lowest BIC was selected.

#### 2 Results of data quality check

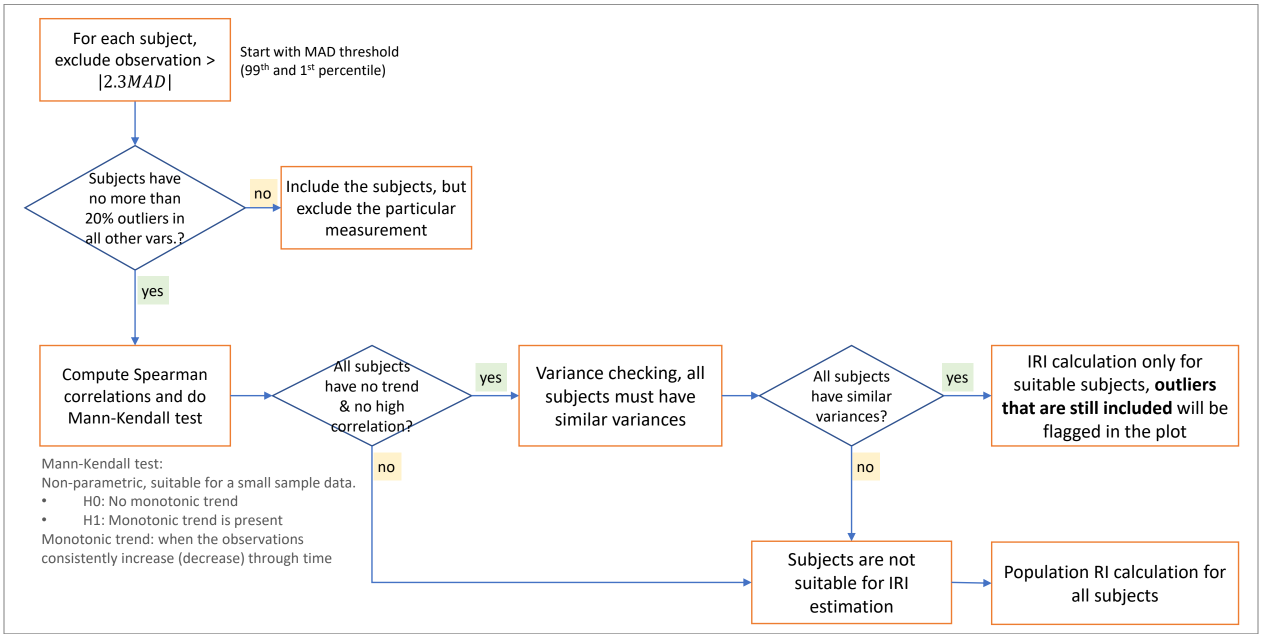


Figure S1: IRI pipeline implemented in the study.

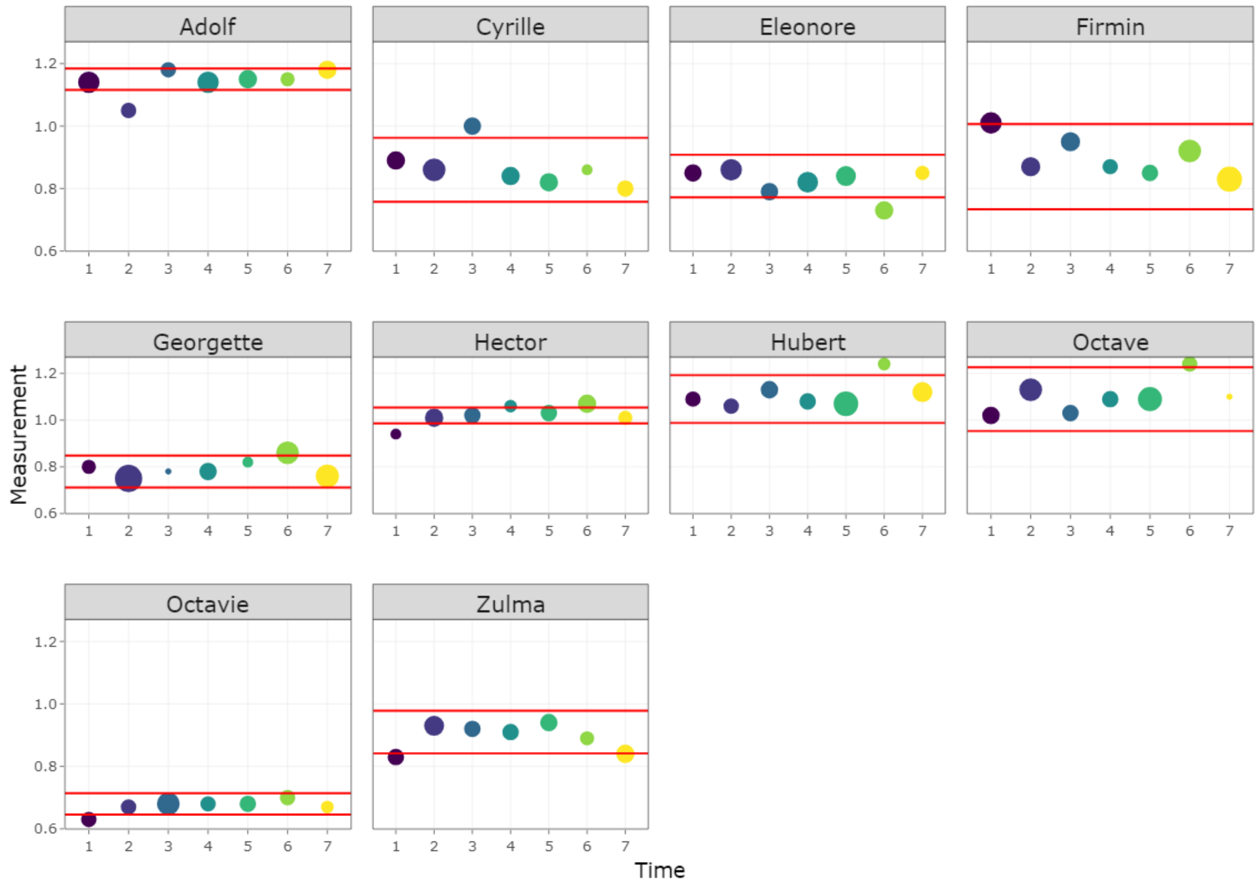


Figure S2: Outlier analysis step of creatinine in the IAF clinical biochemistry data. Outlying observations are shown as the circles outside the MAD thresholds (red horizontal lines).

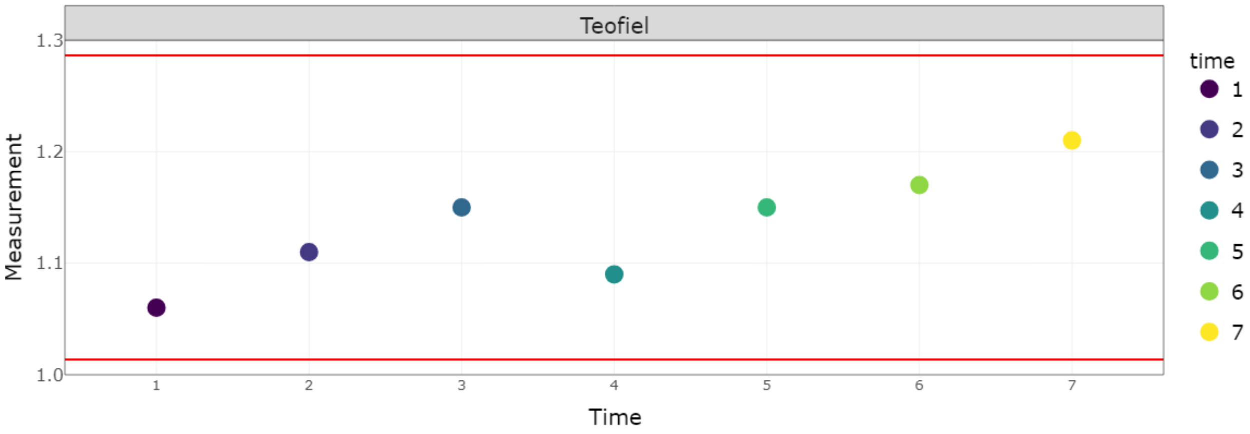


Figure S3: Trend analysis of creatinine in the IAF clinical biochemistry data. This subject has a significant monotonic trend and a high Spearman correlation coefficient ($r\hat{ho} = 0.8649$. This subject will be excluded from the IRI estimation.

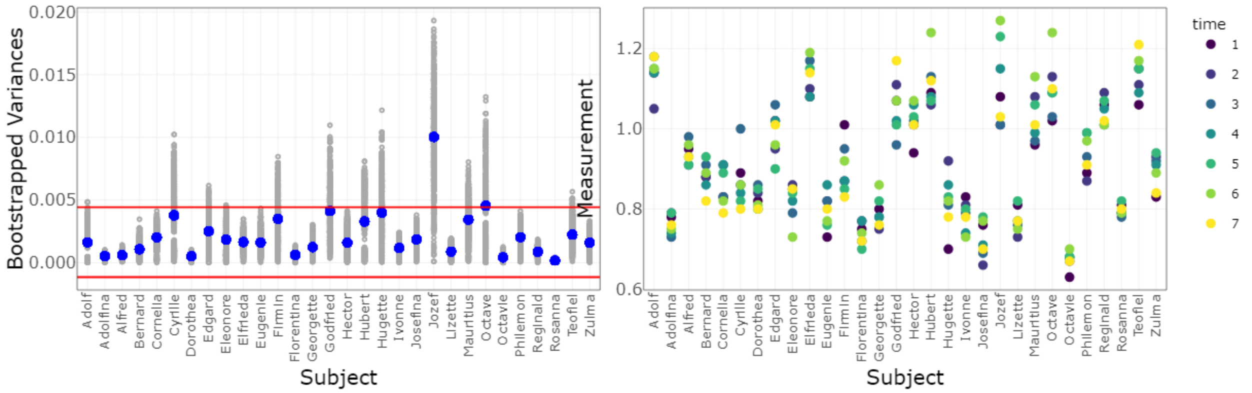


Figure S4: Variance checking of creatinine in the IAF clinical biochemistry data. Two subjects (Jozef and Octave) have variances outside the MAD thresholds and they will be excluded from the IRI estimation.

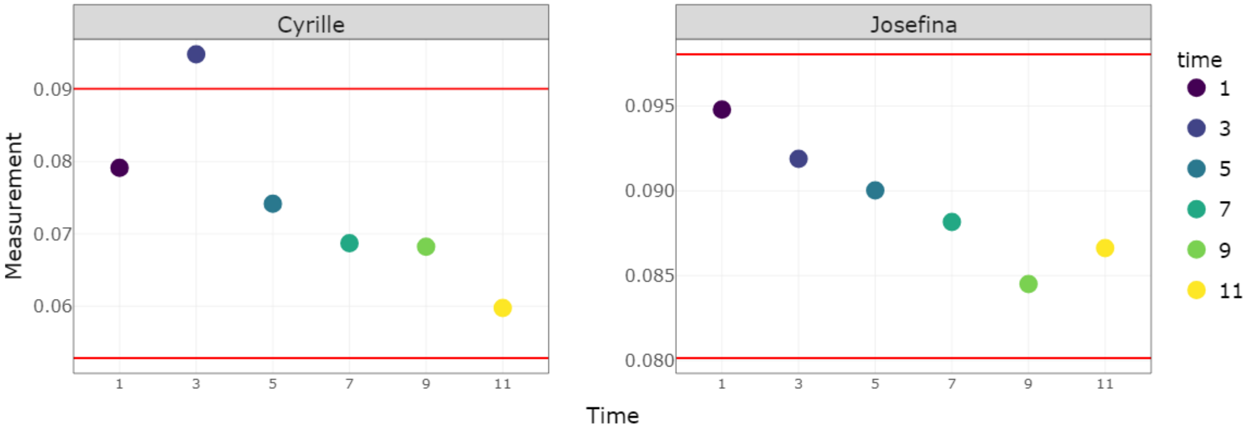


Figure S5: Trend analysis of citrate in the IAF metabolomics data. Two subjects have significant monotonic trends and high Spearman correlation coefficients ($\hat{\rho}_{Cyrille} = -1$ and $\hat{\rho}_{Josefina} = -0.9429$). These subjects will be excluded from the IRI estimation.

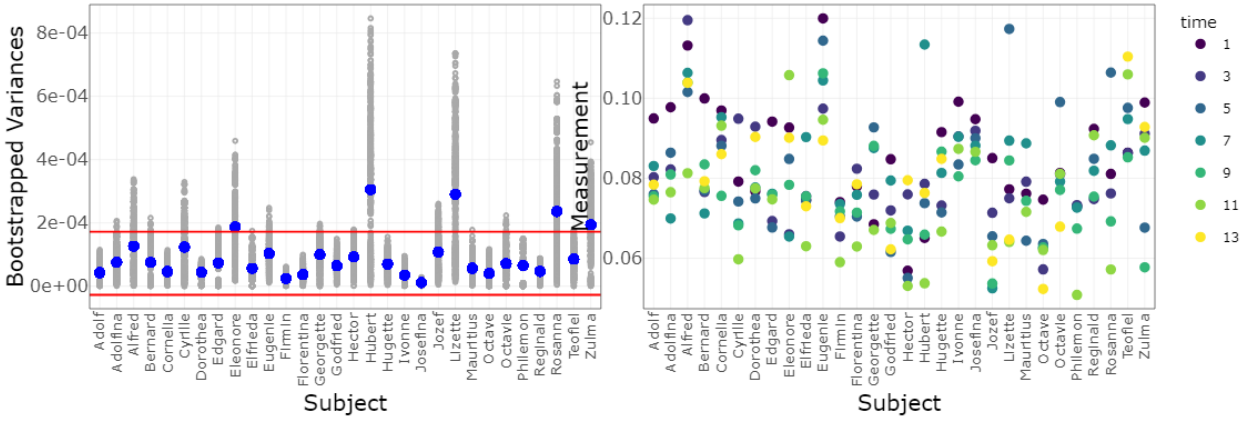


Figure S6: Variance checking of citrate in the IAF metabolomics data. Five subjects have variances outside the MAD thresholds and they will be excluded from the IRI estimation.

### 3 IRI estimates

#### 3.1 Metabolomics and proteomics in the IAF data

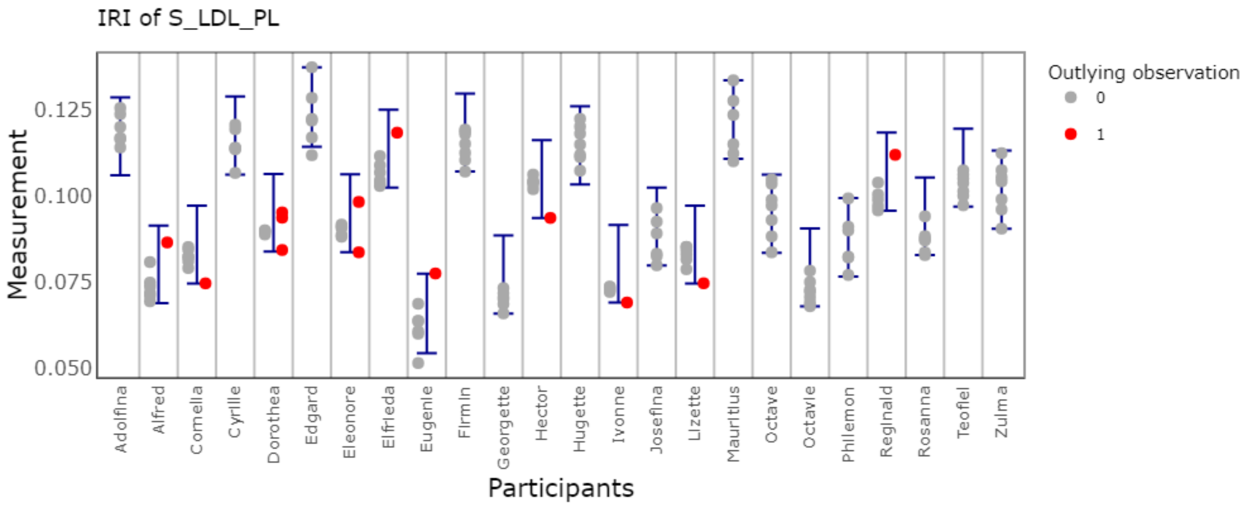


Figure S7: IRI of S-LDL-PL

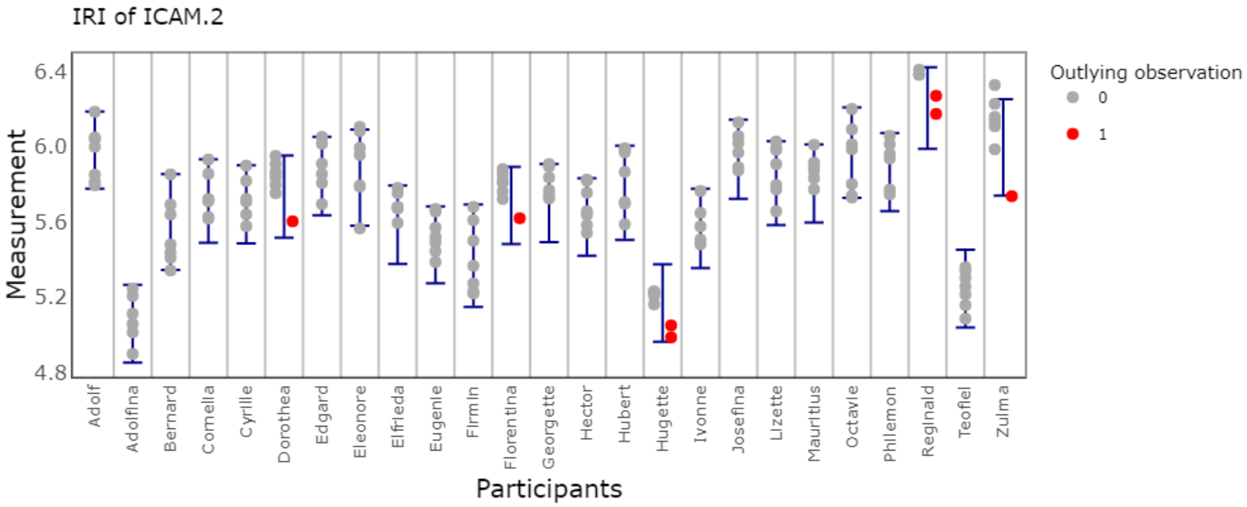


Figure S8: IRI of ICAM-2

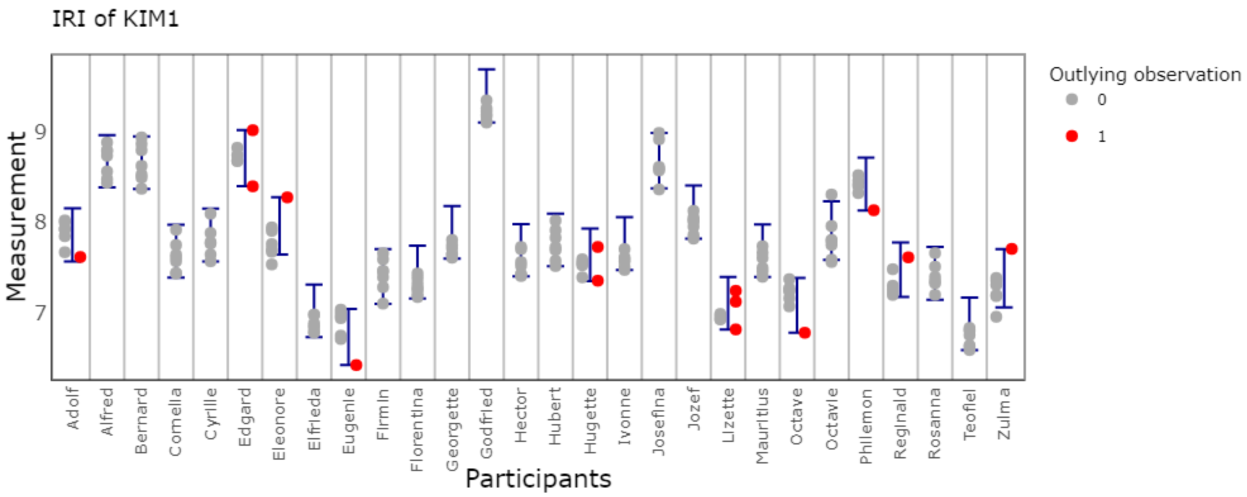


Figure S9: IRI of KIM-1

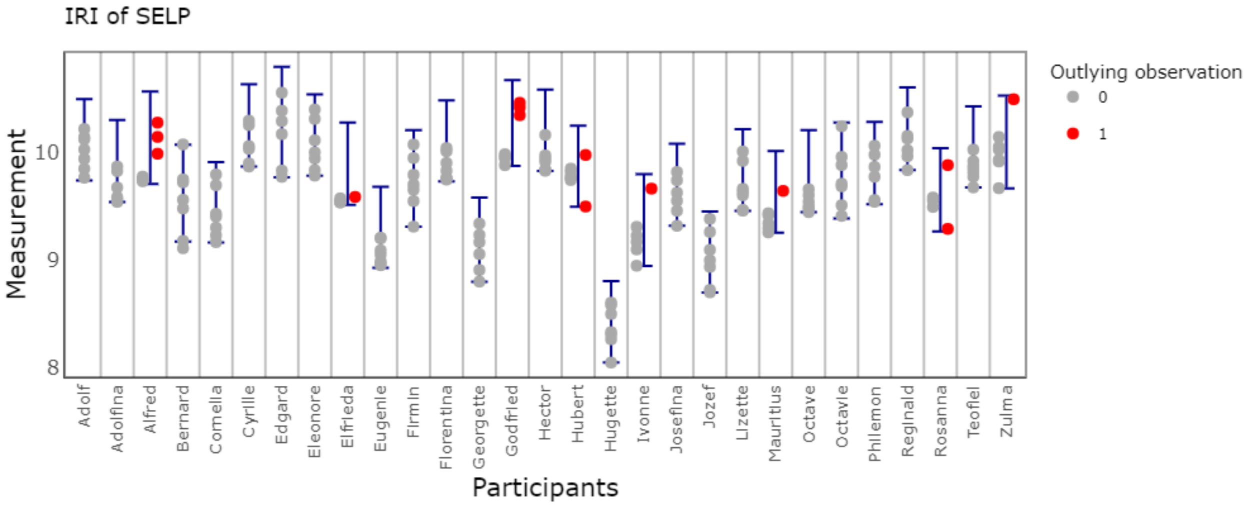


Figure S10: IRI of SELP

#### 3.2 Metabolomics in the IBS data

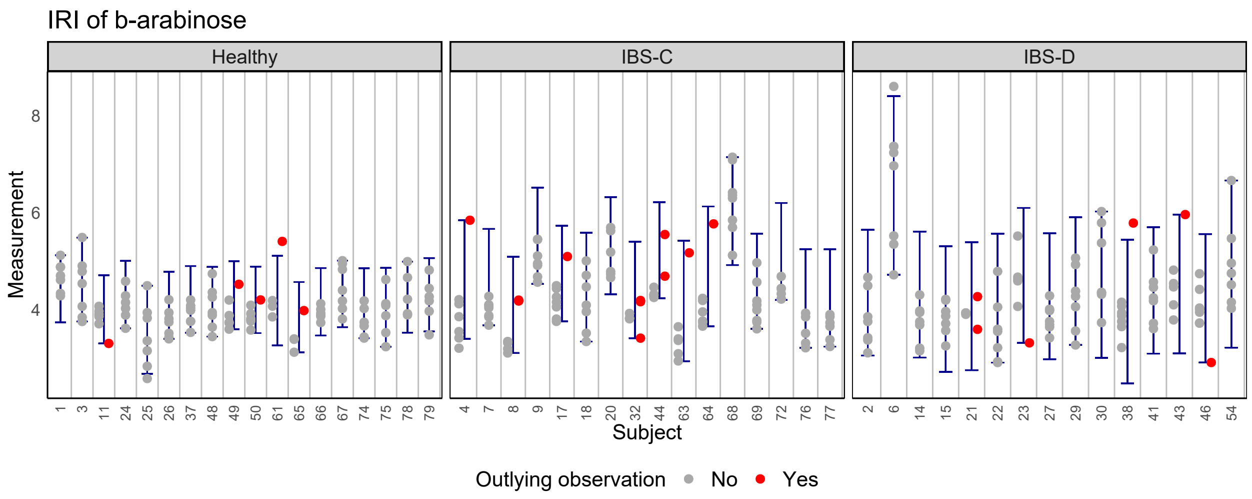


Figure S11: IRI of b-arabinose

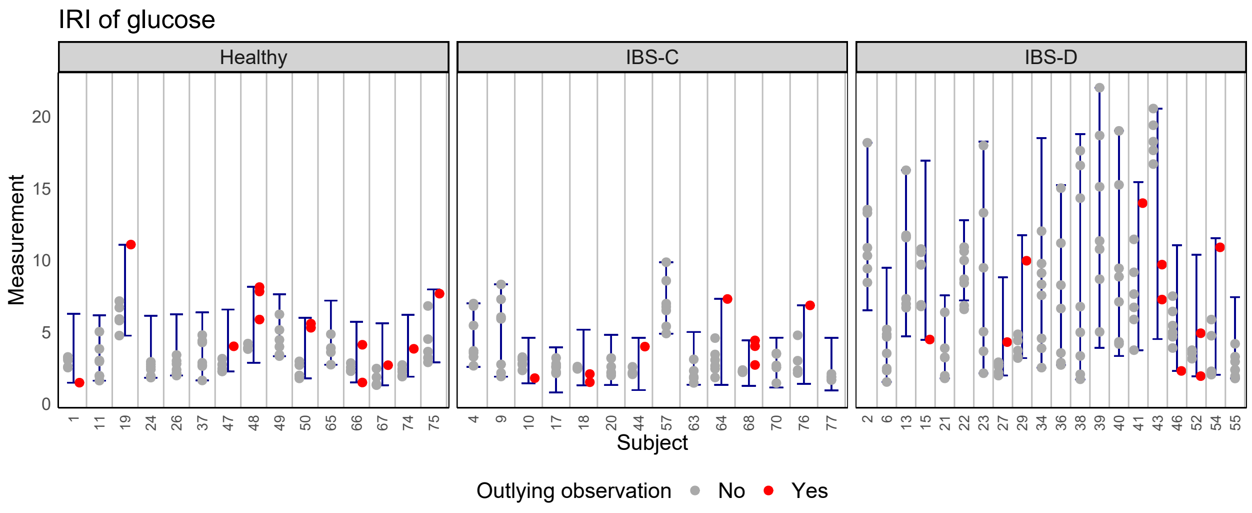


Figure S12: IRI of glucose

Table 1: P-value of the standard student t-test used for comparing the mean of IRI widths between healthy individuals and IBS patients, after the Benjamini-Hochberg correction. The bold font face indicates when the two means are statistically significant at 5% significance level. Metabolites with significant P-values in all three pairs are also made italic.

| Metabolites | Healthy vs IBS-C | Healthy vs IBS-D | IBS-C vs IBS-D |
|---|---:|---:|---:|
| Deoxycholic acid | **0.0455** | 0.0568 | 0.5696 |
| ***Cholic acid*** | **0.0010** | **0.0016** | **0.0007** |
| Ursodeoxycholic acid | 0.0612 | 0.5591 | 0.0628 |
| Lithocholic acid | 0.1373 | **0.0002** | **0.0357** |
| Chenodeoxycholic acid | 0.8482 | **0.0008** | **0.0009** |
| X2 Methylbutyrate | **0.0117** | 0.8693 | **0.0477** |
| ***Lactate*** | **0.0004** | **0.0098** | **0.0000** |
| Alanine | 0.8482 | **0.0058** | **0.0301** |
| Tyrosine | 0.1393 | **0.0005** | **0.0127** |
| Isoleucine | **0.0455** | **0.0002** | 0.1913 |
| Leucine | 0.7971 | 0.1024 | **0.0357** |
| ***Valine*** | **0.0416** | **0.0000** | **0.0026** |
| Lysine | 0.0719 | **0.0130** | 0.2855 |
| Succinate | **0.0455** | 0.0627 | **0.0309** |
| Glycine | 0.8482 | **0.0008** | **0.0007** |
| ***b-arabinose*** | **0.0000** | **0.0000** | **0.0000** |
| ***b-xylose*** | **0.0033** | **0.0000** | **0.0000** |
| Acetate | 0.8457 | 0.0657 | **0.0408** |
| Propionate | **0.0455** | 0.8233 | 0.1927 |
| Butyrate | 0.3211 | 0.2341 | **0.0372** |
| ***Glucose*** | **0.0013** | **0.0000** | **0.0000** |
| Isovalerate | 0.8482 | 0.0752 | 0.0939 |
| Uracil | 0.8482 | 0.2488 | 0.3110 |
| ***Hypoxanthine*** | **0.0000** | **0.0000** | **0.0000** |
